# Validity of using cancer registry data for comparative effectiveness research

**DOI:** 10.1101/19010876

**Authors:** Abhishek Kumar, Zachary D Guss, Patrick T Courtney, Vinit Nalawade, Paige Sheridan, Reith R Sarkar, Matthew P Banegas, Brent S Rose, Ronghui Xu, James D Murphy

## Abstract

**Purpose:** Researchers often analyze cancer registry data to assess for differences in survival among cancer treatments. However, the retrospective non-random design of these analyses raise questions about study validity. The purpose of this study was to determine the extent to which comparative effectiveness analyses using cancer registry data produces results concordant with randomized clinical trials.

**Methods:** We identified 141 randomized clinical trials referenced in the National Comprehensive Cancer Network Clinical Practice Guidelines for 8 common solid tumor types. For each trial we identified subjects from the National Cancer Database (NCDB) matching the eligibility criteria of the randomized trial. With each trial we used three Cox regression models to determine hazard ratios (HRs) for overall survival including univariable, multivariable, and propensity score adjusted models. Multivariable and propensity score analyses controlled for potential confounders including demographic, comorbidity, clinical, treatment, and tumor-related variables. Each NCDB survival analysis was defined as discordant if the HR for the NCDB analysis fell outside the 95% confidence interval of the corresponding randomized trial.

**Results:** NCDB analyses produced HRs for survival discordant with randomized trials in 62 (44%) univariable analyses, 43 (30%) multivariable analyses, and 51 (36%) propensity score models. NCDB analyses produced p-values discordant with randomized trials in 83 (59%) univariable analyses, 76 (54%) multivariable analyses, and 78 (55%) propensity score models. We did not identify any clinical trial characteristic associated with discordance between NCDB analyses and randomized trials including disease site, type of clinical intervention, or severity of cancer.

**Conclusion:** Comparative effectiveness research with cancer registry data often produces survival outcomes discordant to randomized clinical data. These findings help provide context for providers interpreting observational comparative effectiveness research in oncology.

## Introduction

Randomized clinical trials in oncology represent the gold-standard level of evidence from which we establish efficacy of different therapeutic approaches^1^. Despite the critical importance of randomized trials, this study design has acknowledged limitations related to cost, timeliness, and generalizability of results in a “real-world” oncology population^2-4^. Additionally, several clinical scenarios encountered within oncology lack randomized data to support clinical decisions. To help fill these evidence gaps especially in pressing situations, investigators will often rely on research using non-randomized observational data.

Cancer registries within the field of oncology represent a unique resource to study questions surrounding the comparative benefits of different cancer therapies. Cancer registries collect detailed data on incident cancer diagnoses across a substantial portion of the United States (US) population^5-7^. These resources play a critical role in the study of cancer incidence, prevalence, disparities, and patterns of care across the US. However, researchers have increasingly used cancer registry data to evaluate the comparative efficacy of different cancer treatments. The accessibility, ease of use, large sample sizes, and “real world” nature of cancer registry data are attractive for comparative effectiveness research. However, one must also consider potential limitations of using these data for comparative effectiveness research. These include concerns about accuracy with data collection^8^, rigor of analytic techniques^9^, or the potential for selection bias^10^. Together, these limitations could skew results and threaten the validity of comparative effectiveness research with cancer registry data.

Within the field of oncology, we lack a clear understanding of how the survival outcomes of comparative effectiveness research with cancer registry data compare to the gold-standard randomized clinical trials. An improved understanding of this relationship would help providers, researchers, and other consumers of medical literature better assess the role of observational research in the comparative effectiveness landscape. The purpose of this study was to measure agreement (or concordance) in survival estimates between existing randomized clinical trials and comparative effectiveness research with cancer registry data. Furthermore, we sought to identify features of clinical research questions that lead comparative effectiveness research with cancer registry data to more closely align with results of randomized clinical trials.

## Methods

This study identified randomized clinical trials from the literature and compared their survival outcomes to outcomes of patient cohorts systematically analyzed from the National Cancer Database (NCDB). We included common cancer sites including central nervous system, head and neck, breast, lung, gastrointestinal (anal, esophageal, gastric, pancreas, and colorectal cancers), genitourinary (bladder, kidney, and prostate cancers), gynecologic (uterine and cervical cancers), and lymphoma. This study was deemed exempt from institutional review board approval.

### Randomized Clinical Trials

We systematically identified randomized clinical trials referenced in the National Comprehensive Cancer Network (NCCN) Clinical Practice Guidelines for each individual disease site^11-27^. The NCCN Guidelines represent comprehensive tumor site-specific evidence-based consensus-driven guidelines which describe current treatment recommendations. They also include a comprehensive written discussion describing in detail the clinical evidence supporting these recommendations. From the NCCN guidelines we extracted 7,969 referenced publications, and reviewed these references to identify therapeutic randomized clinical trials involving surgery, systemic therapy, hormonal therapy, or radiation therapy. In scenarios where a clinical trial results were reported in more than one publication (typically an “original” publication followed by an “update” at a later date) we kept the most recent publication. **Figure 1** demonstrates our clinical trial selection process, with additional exclusion criteria noted below.

**Figure 1.**
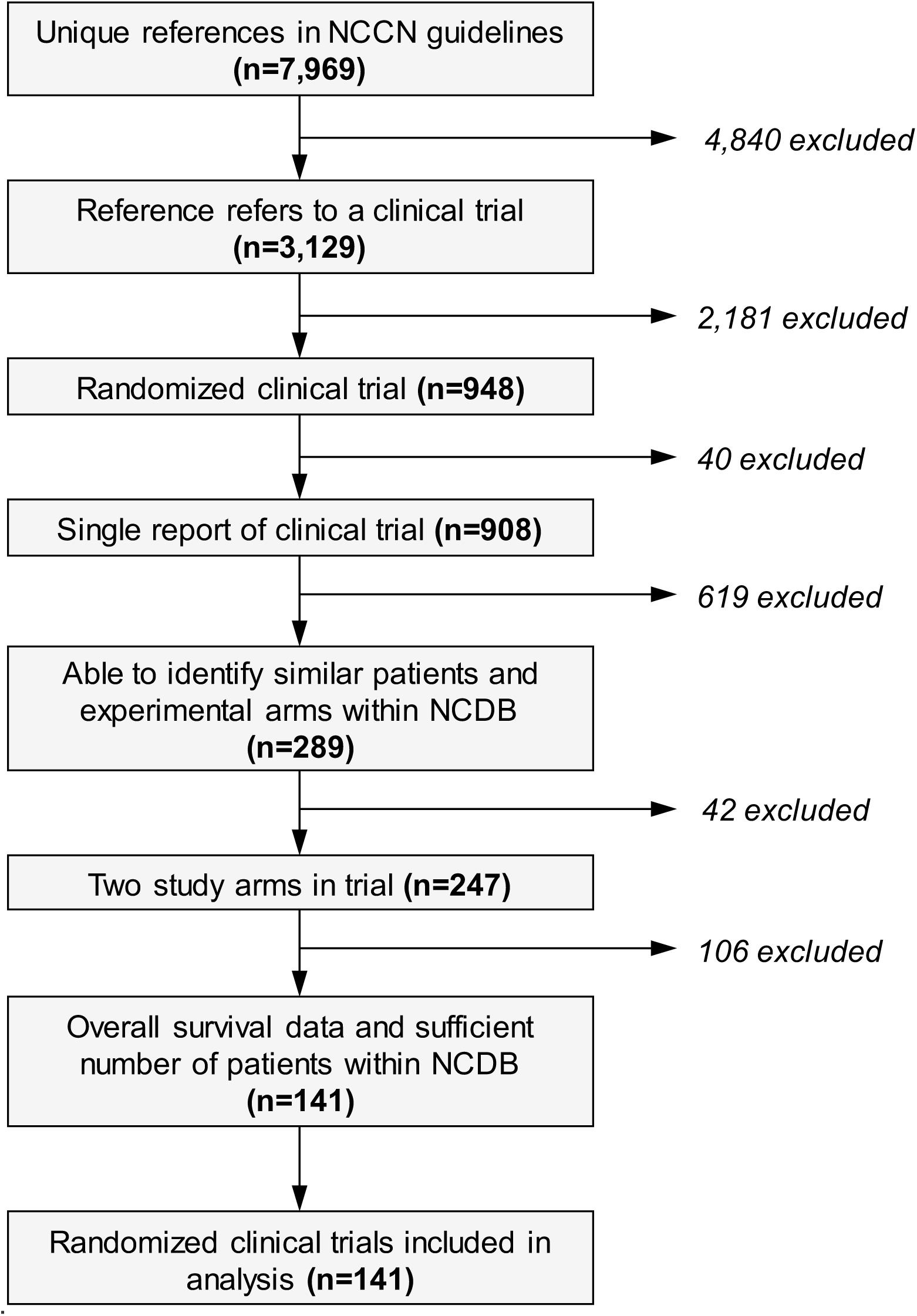
Identification of randomized clinical trials. *Abbreviations:* NCCN = National Cancer Center Network; NCDB = National Cancer Database

### National Cancer Database

The NCDB represents a nationwide facility-based cancer registry sponsored by a collaboration between the American College of Surgeon’s Commission on Cancer and the American Cancer Society. Trained tumor registrars at individual facilities record patient demographic, tumor, and treatment-related information on all incident cancers diagnosed at that facility. Specific information recorded includes tumor, node, metastasis (TNM)-stage, tumor histology, grade, as well as details of treatment including surgery (site, extent of surgery, open vs. laparoscopic) and radiation therapy (dose, number of fractions, treatment modality). Additionally, NCDB records the use of chemotherapy, hormonal therapy, and immunotherapy, though it does not report information on specific agents. Charlson comorbidity score is measured the year prior to diagnosis^28^. This study included patients within NCDB diagnosed between 2004 and 2014.

### Analysis

With each randomized clinical trial we created a cohort of NCDB patients that matched the eligibility criteria of the trial whenever possible. We restricted each NCDB cohort to the disease site in question, histologic confirmation of malignancy, TNM stage per trial eligibility, patient age ≥18 years of age, and no prior history of malignancy. We further restricted each NCDB cohort to select subjects receiving treatments delivered in each clinical trial. For example, if a randomized trial in breast cancer evaluated the impact of mastectomy alone compared to lumpectomy followed by radiation we would include only NCDB patients with a mastectomy alone or those treated with a lumpectomy followed by radiation therapy. Due to a lack of available data on specific systemic agents, we were unable to include clinical trials comparing single-agent systemic regimens to multi-agent systemic regimens, or clinical trials evaluating two different systemic agents. We did include randomized trials where one arm contained a systemic agent and the other arm did not, with the limitation that the specific agent used was unknown. Randomized clinical trials often have eligibility criteria which include patient performance status or baseline laboratory values. Similar to other cancer registries, NCDB does not contain performance status or laboratory data, therefore these factors were not considered in our analysis. For trials involving radiation therapy, we restricted radiation doses to within 10 Gy of the trial specified dose. If the trial specifically compared two different radiation doses, we restricted the NCDB cohort to the specified dose and number of fractions delivered in the trial. Within the NCDB cohorts we defined chemotherapy delivered concurrently with radiation if the start dates for both modalities occurred within 14 days of each other.

### Statistical Analysis

Of the randomized clinical trials included in this analysis we recorded the hazard ratio for overall survival along with the 95% confidence interval and associated p-value. For trials that did not present a hazard ratio for overall survival in the manuscript we estimated the hazard ratios from Kaplan-Meier plots^29^. With each cohort of NCDB patients, we determined hazard ratio for overall survival using three different analytic approaches including: 1) unadjusted univariable Cox proportion hazards model; 2) multivariable Cox proportional hazards model; and 3) propensity score adjusted analysis. All models measured survival from the date of diagnosis through death of any cause, censoring at the date of last follow-up. We conducted a sensitivity analysis measuring survival from the date of treatment initiation, which did not influence our results (analysis not presented). Multivariable models included known confounding variables available in NCDB which could potentially influence survival. At a minimum each multivariable model included patient age, gender, race, geographic region, median household income, Charlson Comorbidity score, year of diagnosis, grade, site-specific histology, and clinical or pathologic TNM stage. Individual multivariable models included additional factors related to specific disease sites, including ER/PR/HER2+ status for breast cancer, as well as prostate-specific antigen (PSA) and Gleason score for prostate cancer. With the propensity score analysis for each NCDB cohort, we derived propensity scores from logistic regression models using the same covariates noted above in the multivariable analyses. With each propensity score analysis we used inverse probability of treatment weighting, and assessed balance between weighted covariates with standardized differences.

We assessed concordance between the randomized clinical trials and corresponding NCDB analyses using three different approaches. First, we assessed the relationship between hazard ratios for overall survival between the clinical trial and NCDB analysis with a Pearson correlation coefficient, with a score of 0 indicating no correlation, and 1 indicating perfect correlation. Second, we assessed concordance in hazard ratios with an NCDB analysis considered *concordant* if the NDCB hazard ratio fell within the 95% confidence interval of the hazard ratio from the clinical trial^30^. Third, we considered concordance with respect to statistical significance, with an NCDB analysis considered *concordant* if both the NCDB and clinical trial p-values for survival were non-significant (p>0.05), or if they were both significant (p<0.05) with survival favoring the same treatment arm in NCDB and the clinical trial.

Univariable logistic regression was used to identify whether certain features of a clinical trial would lead to higher levels of concordance in hazard ratios between the clinical trial and NCDB analysis. We analyzed the following features: domestic versus international trial, year of trial publication, tumor site, type of clinical trial question addressed, difference in mean age between NCDB treatment arms, and aggressiveness of cancer (assessed with the proxy of median survival of study population). We also addressed the impact of trials with treatment arms of different durations to indirectly address the influence of immortal time bias^31^. A study was defined susceptible to immortal time bias if one treatment arm included lengthier treatment than the other treatment arm. Statistical analyses were performed using SAS version 9.4 (SAS Institute, Cary, NC), with p-values less than 0.05 considered statistically significant.

## Results

One hundred and forty-one randomized clinical trials met inclusion criteria (**Figure 1**), with characteristics of the studies provided in **Table 1**, and a complete list of trial references provided in **Supplemental Table 1**. In general, the majority of clinical trials were published between 2004 and 2014, took place outside North America, and the trials addressed an array of clinical questions though nearly half evaluated the utility of adding systemic therapy to surgery or radiation. In total the clinical trials included 85,118 patients whereas the corresponding NCDB analyses included 1,344,536 patients. The median size of a clinical trial was smaller than the median size of an NCDB analysis (396 patients vs. 6,543 patients).

**Table 1.** Characteristics of randomized clinical trials

| **Characteristic** | **No. (%)** |
| --- | --- |
| **Year of trial publication** |  |
| 2003 or earlier | 31 (22) |
| 2004-2014 | 92 (65) |
| 2014 or later | 18 (13) |
| **Geographic region of trial** |  |
| North America | 43 (31) |
| Outside North America | 88 (62) |
| Both | 10 (7) |
| **Anatomic site of tumors** |  |
| Breast | 19 (13) |
| Central nervous system | 8 (6) |
| Gastrointestinal | 47 (33) |
| Genitourinary | 18 (13) |
| Gynecologic | 13 (9) |
| Head and neck | 14 (10) |
| Lymphoma | 6 (4) |
| Lung | 16 (11) |
| **Research question** |  |
| Addition of systemic therapy | 67 (48) |
| Addition of radiation | 18 (13) |
| Addition of radiation and systemic therapy | 11 (8) |
| Addition of surgery | 6 (4) |
| Radiation dose | 11 (8) |
| Timing of treatment | 5 (4) |
| Type of surgery | 6 (4) |
| Type of systemic therapy | 7 (5) |
| Other | 10 (7) |

When considering all clinical trials, we found a weak correlation between the hazard ratios (HRs) from the clinical trials and the HRs from the NCDB analysis for all three statistical approaches (**Figure 2**). The correlation between the HR from the clinical trials and NCDB was weakest with the unadjusted analysis (r=0.20; 95% confidence interval [CI]=0.03-0.35, p=0.02), followed by the propensity score analysis (r=0.25; 95% CI=0.09-0.40, p=0.003), and followed by the multivariable analysis (r=0.34, 95% CI=0.18-0.48, p<0.001).

**Figure 2.**
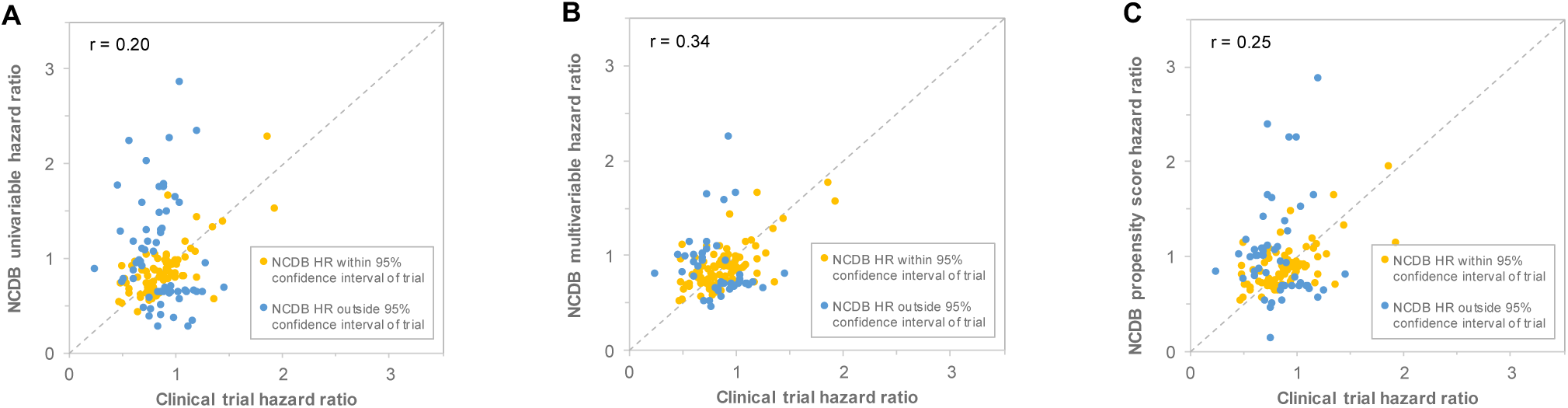
Comparison of hazard ratios from randomized clinical trials and analyses with data from NCDB. Each point on the scatter plot represents the hazard ratio for overall survival from one of the 141 randomized clinical trials in this study and the corresponding analysis within NCDB. Yellow dots represent NCDB analyses where the NCDB hazard ratio falls within the 95% confidence interval of the hazard ratio in the clinical trial, whereas blue dots represent hazard ratios from the NCDB analyses that fall outside the 95% confidence interval of the hazard ratio in the clinical trial. The grey dashed line represents the line where clinical trial hazard ratios equal NCDB hazard ratios. The individual panels represent univariable analyses (A), multivariable analyses (B), and propensity score analyses (C).

When evaluating concordance in hazard ratios for overall survival we found that 62 of 141 trials (44%) evaluated in NCDB with a univariable analysis produced hazard ratios discordant with compared to clinical trials. With multivariable analysis, 43 trials (30%) analyzed in NCDB produced hazard ratios discordant with randomized trials. With propensity score analysis, 51 trials (36%) analyzed in NCDB produced hazard ratios discordant with randomized trials. **Supplementary Figure 1** demonstrates results of all clinical trials in the analysis.

When evaluating discordance with statistical significance we found that 83 trials (59%) analyzed with a univariable analysis, 76 trials (54%) analyzed with a multivariable analysis, and 78 trials (55%) analyzed with a propensity score analysis produced p-values discordant with clinical trials (**Table 2**). Forty-nine clinical trials found one treatment arm to have significantly improved survival, yet NCDB analysis found discordant p-values in 21 trials (43%) analyzed with a univariable analysis, 19 trials (39%) analyzed with a multivariable analysis, and 19 trials (39%) analyzed with a propensity score analysis. Ninety-two clinical trials found no significant difference in overall survival between treatment arms, yet NCDB found significant p-values in 62 trials (67%) analyzed with univariable analysis, 57 trials (62%) analyzed with a multivariable analysis, and 59 trials (64%) analyzed with a propensity score analysis.

**Table 2.**
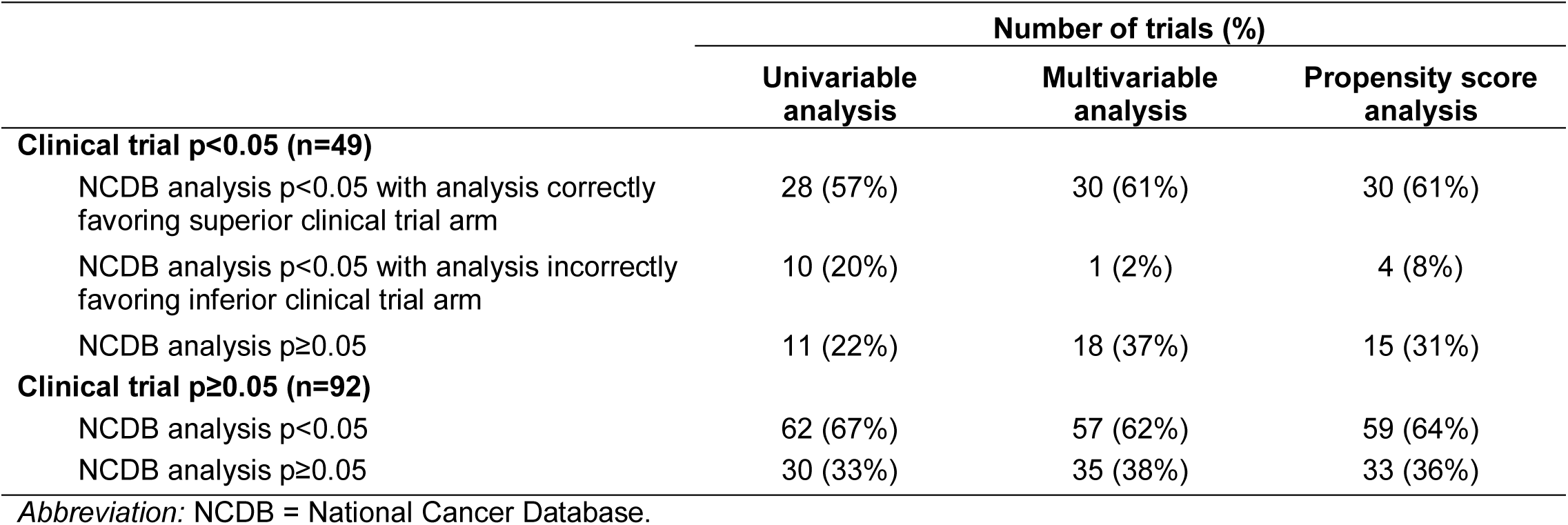
Concordance in statistical significance between clinical trials and NCDB analysis

Analysis to identify individual predictors of discordance did not find any factor associated with discordance in hazard ratios for overall survival between NCDB and clinical trials (**Figure 3**).

**Figure 3.**
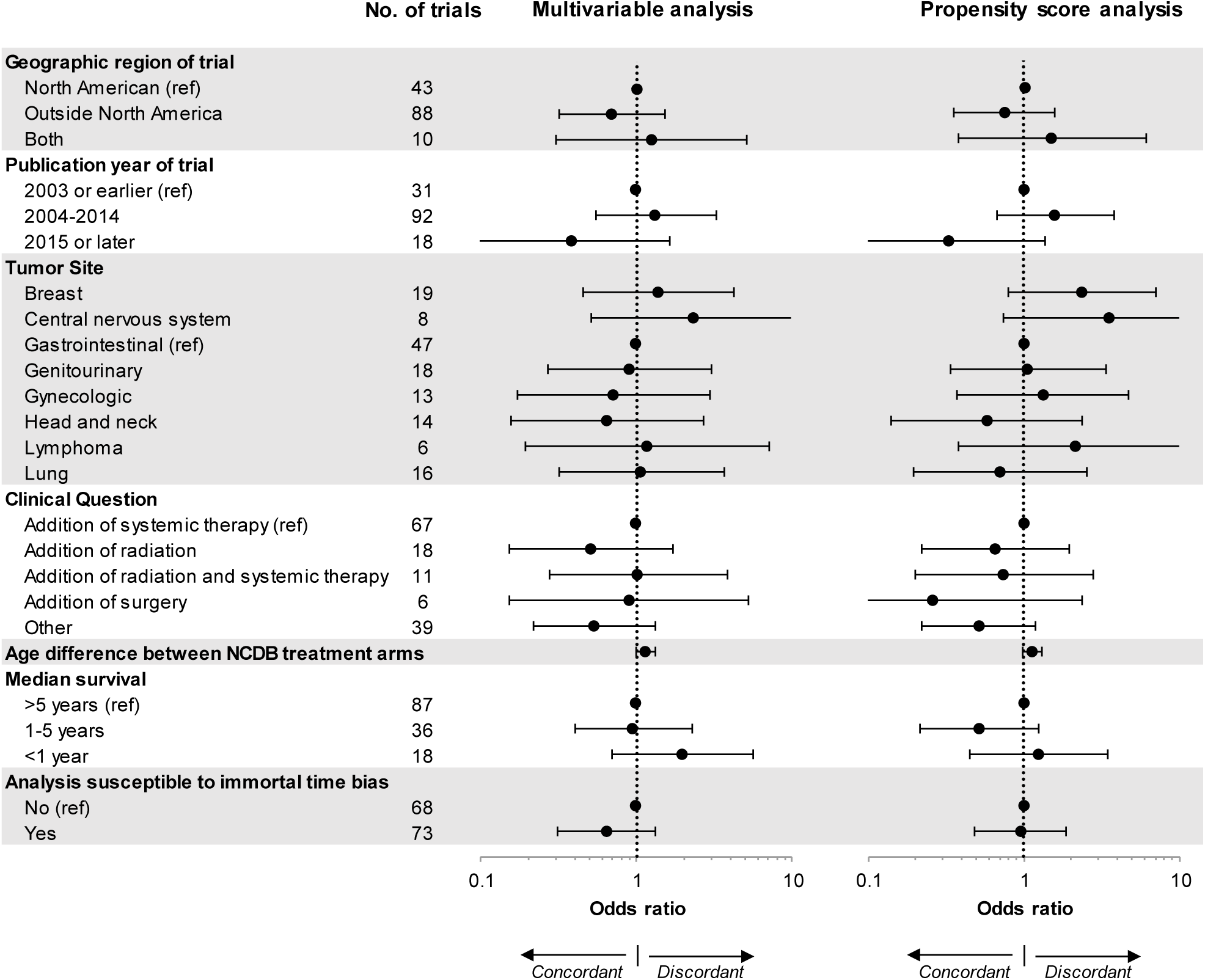
Predictors of discordance in hazard ratios with clinical trials and NCDB. This forest plot demonstrates univariable logistic regression analyses to identify factors that predict when an NCDB analyses will produce results discordant with randomized clinical trials. Discordance NCDB analyses were defined as occasions when the NCDB hazard ratio falls outside the 95% confidence interval of the hazard ratio in the clinical trial. Odds ratios greater than 1 indicate NCDB analyses were more likely to be discordant with clinical trials. *Abbreviations:* NCDB = National Cancer Database; ref = reference.

## Discussion

This study demonstrates substantial discordance and little correlation between survival outcomes of randomized clinical trials and comparative effectiveness research using cancer registry data. Multivariable models and propensity score analyses improved concordance modestly, yet approximately one-third of clinical trials replicated with cancer registry data produced overall survival outcomes discordant with clinical trials. Furthermore, this study did not identify factors associated with the clinical research question which might help identify cancer registry research studies more or less in line with the results of randomized clinical trials.

These findings complement other research in oncology^30^ and non-oncology fields^32-34^ which evaluates differences in outcomes between randomized clinical trials and observational research. Within oncology, Soni et al. recently published a study using different methodology comparing the results of published comparative effectiveness research to published randomized clinical trials^30^. Soni found that 38% of analyses with cancer registry data produced survival results discordant with clinical trial data, whereas our study found discordance rates between 30-44% depending on the analytic method. Our study methodology differed from Soni in that we used a standardized and consistent approach to analyze the cancer registry data as opposed to extracting results from the literature which used more heterogeneous analytic approaches. Regardless, these analyses together suggest that a substantial portion of comparative effectiveness research using cancer registry data may produce results that differ substantially from randomized clinical trials.

Our analysis of statistical significance found discordance between observational cancer registry research and clinical trial outcomes, though one must consider that calculations of statistical significance depends exquisitely on sample size. Therefore, the larger number of patients within cancer registries inherently leads to a higher likelihood of a lower p-value. Despite this issue, one must also consider that physicians frequently misunderstand the basic concepts of p-values^35,36^. Furthermore, the overall utility of the p-value in research represents an active subject of debate^37^, though one must accept that treatment and policy recommendations will often depend on the question of statistical significance^38-40^.

Multiple factors could potentially explain the observed differences between comparative effectiveness research with registry data and randomized clinical trials, though selection bias represents a key factor worth discussing. With observational research the lack of randomization raises the important possibility of *selection bias* which represents an imbalance in known or unknown confounding factors between treatment groups. Factors such as body mass index, tobacco use or performance status – not recorded in cancer registry data – could influence treatment decisions and independently impact survival regardless of the treatment chosen. One could hypothesize that more granular databases, such as cancer data linked to electronic health record data, could help better control for these confounding factors, though the magnitude of how much these factors influence results remains an open question which deserves further study.

Beyond selection bias, one must consider the potential influence of misclassified cancer registry data. The Charlson index estimates a patient’s comorbidity, though research suggests that this index may provide only a rough measure of underlying comorbidity^41^, and additional research suggests that Charlson comorbidity may be underreported within cancer registries^42^. Similarly, research demonstrates the potential for underreporting of systemic therapy, hormonal therapy, and radiation therapy in cancer registry data^8^. Overall, we lack a clear understanding of whether these misclassified variables occur at random which would tend to bias results towards the null (no survival difference), or occur not at random which could have an unpredictable influence on survival analyses.

Beyond the aforementioned issues one must consider that even with perfect cancer registry data the outcomes between observational cancer registry research and randomized trials might fundamentally differ. Patients in clinical trials represent a younger group of patients with better performance status and fewer comorbidities compared to the general oncology population^43^. Differences in outcomes between clinical trials and comparative effectiveness research with registry data could potentially reflect a differential effect of treatment in healthier clinical trial patients^44^. Similarly, one must consider the possibility that the technical delivery of cancer therapy in the “real world” may differ from delivering cancer therapy under the constraints of a clinical trial. In this case, one could potentially consider differences in observational data as a potential “implementation failure” as opposed to a true “treatment failure”. Overall, it is difficult to identify *one* reason accounting for the differences in outcomes between observational research and clinical trials. Ultimately these differences in outcomes likely stem from multiple contributing factors.

This current study has limitations worth discussing. This study focused on analyzing discordance in overall survival and could not evaluate cancer-specific survival due to the lack of cause of death information within NCDB. While overall survival represents a common primary endpoint in randomized clinical trials, one could hypothesize that cancer-specific survival might represent an endpoint more likely to produce concordant results between observational research studies and clinical trials. This study focused on data within NCDB because of the detailed treatment information provided with this dataset. We cannot comment on whether the observed findings in this study hold with other observational datasets including SEER^7^, administrative datasets, or more granular institution-specific datasets. This study attempted to recreate clinical trials with cancer registry data, though we could not account for all inclusion/exclusion criteria associated with clinical trials including chemotherapy details (agents, dose and schedule), hormone therapy agents, radiation techniques, and specific details of surgery. This limitation holds true with this current analysis as well as any research involving cancer registry data. Finally, this study used a structured approach to analyzing cancer registry data, though one must acknowledge that incorporating different variables into our multivariable models, or employing different propensity score analytic techniques could potentially influence these outcomes.

Despite these limitations this study demonstrates substantial discordance in survival outcomes between comparative effectiveness research with cancer registry data and randomized clinical trial outcomes. These discrepant findings could represent fundamental differences in patient selection or treatments rendered in clinical trials compared to the real world. Alternatively, they could stem from issues surrounding selection bias or misclassification of data within cancer registries. Regardless of the underlying cause, understanding these differences will help provide context to providers and researchers as they interpret comparative effectiveness research in oncology.

## Data Availability

All data referred to in the manuscript is publicly available through the National Cancer Database (NCDB).

